# Improved detection of aberrant splicing using the Intron Jaccard Index

**DOI:** 10.1101/2023.03.31.23287997

**Authors:** Ines F. Scheller, Karoline Lutz, Christian Mertes, Vicente A. Yépez, Julien Gagneur

## Abstract

Detection of aberrantly spliced genes is an important step in RNA-seq-based rare disease diagnostics. We recently developed FRASER, a denoising autoencoder-based method for aberrant splicing detection that outperformed alternative approaches. However, as FRASER’s three splice metrics are partially redundant and tend to be sensitive to sequencing depth, we introduce here a more robust intron excision metric, the Intron Jaccard Index, that combines alternative donor, alternative acceptor, and intron retention signal into a single value. Moreover, we optimized model parameters and filter cutoffs using candidate rare splice-disrupting variants as independent evidence. On 16,213 GTEx samples, our improved algorithm called typically 10 times fewer splicing outliers while increasing the proportion of candidate rare splice-disrupting variants by 10 fold and substantially decreasing the effect of sequencing depth on the number of reported outliers. Application on 303 rare disease samples confirmed the reduction fold-change of the number of outlier calls for a slight loss of sensitivity (only 2 out of 22 previously identified pathogenic splicing cases not recovered). Altogether, these methodological improvements contribute to more effective RNA-seq-based rare diagnostics by a drastic reduction of the amount of splicing outlier calls per sample at minimal loss of sensitivity.

## INTRODUCTION

The regulation of splicing is important to control isoform expression and cellular function (1–3). Defects at the level of the pre-mRNA splicing process represent a major cause of human disease. It is estimated that 15–50% of all variants leading to disease in humans alter splicing (4, 5). Different methods that predict the impact of a variant in splicing from sequence alone have been developed (6–12). However, even with increasing precision, their accuracy remains imperfect, especially for diagnostics and for variants located far from the splice sites (9, 12). Even the ACMG guidelines for variant pathogenicity require additional functional evidence such as RNA-seq for variants predicted to disrupt splicing (13). Importantly, current prediction models do not inform on the consequence of a potential splicing defect on the resulting transcript isoform (e.g. frameshift or exon truncation). Identifying splicing aberrations on RNA sequencing (RNA-seq) data provides more direct evidence of the presence of splicing defects and reveals the resulting transcript isoforms. This approach has been successfully used to diagnose patients with rare genetic disorders in large cohorts (14–20). Following this initial success, computational tools specialized on detecting aberrant splicing from RNA-seq data have been developed including LeafcutterMD (21), SPOT (22), and FRASER (23). FRASER (23) models three metrics that are computed from split reads and unsplit reads detected at de novo identified splice sites. These three metrics are the percent-spliced-in to test the splice acceptor site (*ψ*_5_) and donor site (*ψ*_3_), and splicing efficiency (*θ*). These metrics capture different types of aberrant splicing, namely aberrant acceptor site usage, aberrant donor site usage and aberrant splicing efficiency. FRASER uses a denoising-autoencoder approach to control for potentially unknown sources of covariation between samples, and calculates beta-binomial *P*-values to identify splicing outliers. Benchmarks on rare variant enrichment among reported splicing outliers showed that FRASER outperformed LeafcutterMD and SPOT.

Despite FRASER’s improvements, the number of outlier calls per sample often remains very large. Notably, these splice-site-centric metrics can lead to reporting outliers that reflect local aberrations that may only have minor effects on the abundance of the canonical splicing isoform. Figure 1A illustrates such a case, where an exon elongation appearing in one sample is supported by 1,017 reads, while there are 32,018 supporting the annotated intron. Taking as reference the newly created acceptor site, this exon elongation which is present in only 6 out of 582 samples, appears as an outlier event. Consistently, FRASER detected this event significantly and with a differential *ψ*_5_ = 0.85. However, this event does not strongly affect the canonical isoform distribution because the vast majority of the reads still support the annotated intron.

**Figure 1.**
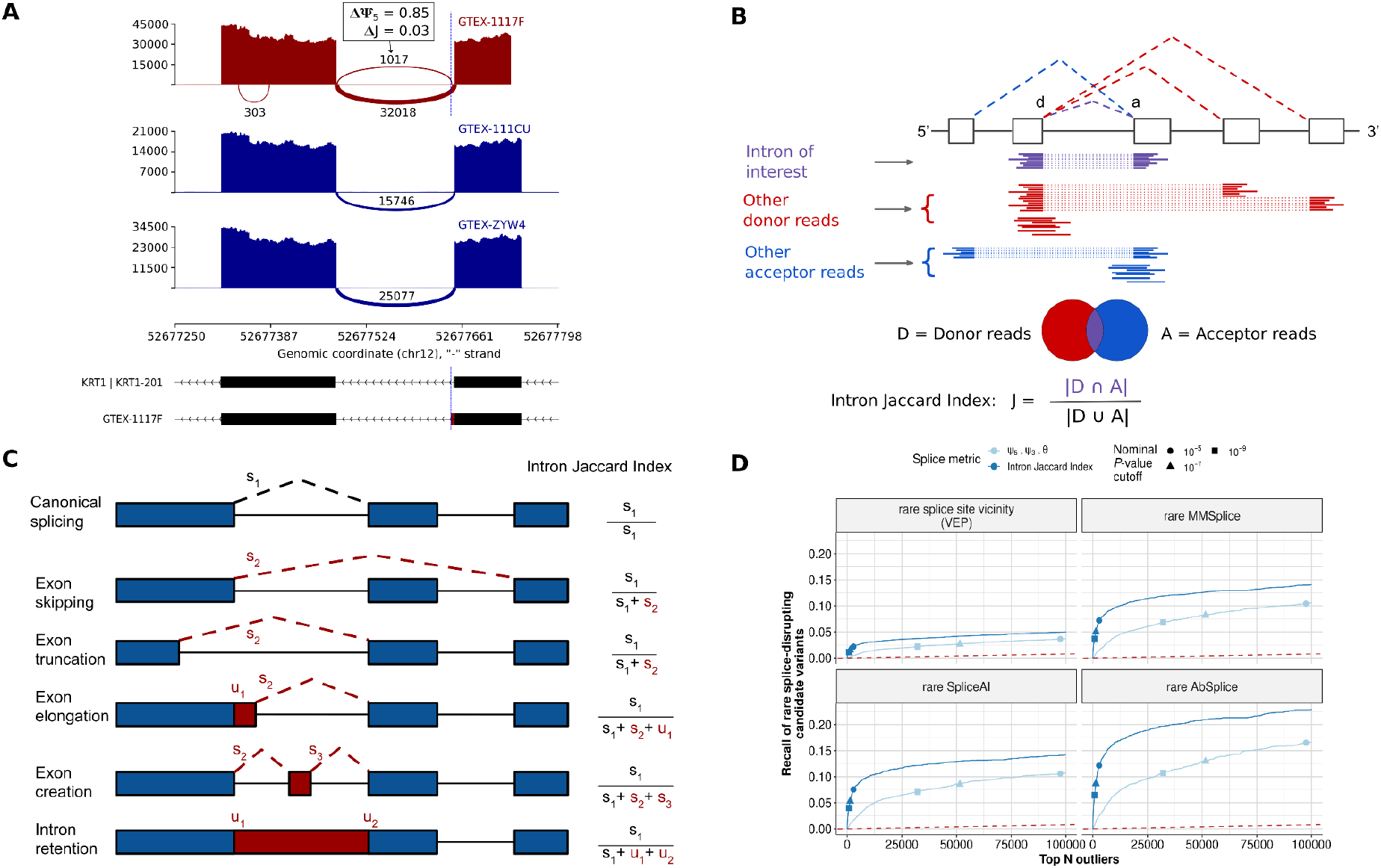
Intron Jaccard Index metric improves splicing outlier calling. **(A)** Sashimi plot of three GTEx skin not-sun-exposed RNA-seq samples showing exons 4 and 5 of the *KRT1* gene. A splicing outlier was detected in the top sample using the *ψ*_5_ metric of FRASER (red). The position of the donor site of the outlier intron is indicated with a blue dashed line. While this intron is not expressed in most other samples (dark blue) and therefore detected with a high Δ*ψ*_5_ value, its functional impact is probably minor because the canonical intron remains largely dominant. **(B)** Schematic definition of the Intron Jaccard Index metric. **(C)** Representation of different types of aberrant splicing events that can be captured with the Intron Jaccard Index metric. The right column contains the formulae to compute the Intron Jaccard Index metric of the canonical intron (black dotted line) from the split (*s*) and non-split (*u*) reads of the involved introns in each scenario. **(D)** Recall of rare splice-disrupting candidate variants (as defined by VEP, MMSplice, SpliceAI and Absplice) versus the rank of nominal *P*-values from FRASER (light blue) and from an adaptation of FRASER using the Intron Jaccard Index metric (dark blue) on the GTEx skin not-sun-exposed dataset (*N*=582). Different nominal *P*-value cutoffs are indicated with shapes.

To address this issue, we here introduce a new intron-based metric which we named the Intron Jaccard Index. It is defined as the proportion of reads supporting the splicing of an intron of interest among all reads associated with either splice site of the intron (Figure 1B). The Intron Jaccard Index is computed using both split and non-split reads, thus allowing to capture several types of aberrant splicing, including exon skipping, exon truncation, exon elongation, exon creation and full intron retention (Figure 1C). The Inton Jaccard Index is conceptually similar to the intron excision ratio concept underpinning LeafCutter (24). However, the statistical model of LeafCutter models multiple introns of a same locus jointly, which leads to modeling complications and does not allow for modeling sample co-variation. Here we model the Intron Jaccard Index of individual introns separately, using the same beta-binomial autoencoder approach as in FRASER. We furthermore perform a systematic evaluation of model parameters including pseudocounts and filtering criteria. Collectively we refer to this new method as FRASER 2.0. We next benchmark FRASER 2.0 against FRASER, LeafCutterMD, and SPOT using the multi-tissue GTEx dataset. Finally, we apply FRASER 2.0 to independent rare disease cohorts and validate it on previously reported pathogenic splice defects.

## MATERIAL AND METHODS

### Datasets

The GTEx dataset consists of 17,350 RNA-seq samples from 54 tissues of 948 assumed healthy individuals of the Genotype-Tissue Expression Project V8 (25). The GTEx data used for the analyses described in this manuscript were obtained from dbGaP accession number phs000424.v8.p1. Samples with a RIN number < 5.7 and tissues with less than 100 samples were discarded. This resulted in a total of 16,146 samples and 48 tissues.

The Yépez et al. dataset consists of 303 individuals affected with a rare mitochondrial disorder described in (20). The intron counts were downloaded from Zenodo (26, 27).

The Undiagnosed Disease Network (UDN) (28) dataset consists of individuals suffering from a rare genetic disorder, as well as unaffected controls collected from different centers in the United States. We downloaded it from dbGaP (study accession phs001232.v4.p2). It contains 821 RNA-seq samples extracted from blood (*N*=370), fibroblasts (*N*=398), and other tissues (*N*=53) which were not further considered. All RNA-seq samples are stranded except for 1 which was removed. Poly-A sequenced samples with a high-quality exonic rate lower than 0.7 were removed. Samples with size factors, a sequencing depth metric (29), larger than 3 or lesser than 0.25 were also discarded. The resulting dataset consists of 252 poly(A) blood, 104 total RNA blood, and 391 fibroblast samples.

### Splicing outlier detection with FRASER

The integrated workflow DROP v1.1.3 (30) was used to count split and non-split reads from the BAM files (from the GTEx and UDN datasets) and to detect splicing outliers with FRASER v1.2.2 (23) on all datasets. Default cutoffs (FDR < 0.1, |Δ*ψ*| ≥ 0.3, minimal intron coverage ≥ 5 reads) and default intron filtering settings (95% of samples with N ≥ 1 and at least one sample with an intron count ≥ 20) were used.

### Splicing quantification using the Intron Jaccard Index metric

We defined the Intron Jaccard Index (J) as the Jaccard index of the sets of donor-associated reads *D* and acceptor-associated reads *A*. For a given sample *i* and intron *j*, the set of donor associated reads *D*_*ij*_ was defined as the set of all split-reads with the same donor site as intron *j* as well as all non-split reads spanning the exon-intron boundary at that donor site of sample *i*. The set of acceptor-associated reads *A*_*ij*_ was analogously defined for the split and non-spit reads at the acceptor site of intron *j* (Figure 1B).

The Intron Jaccard Index *J*_*ij*_ for sample *i* and intron *j* w*a*s calculated as follows:

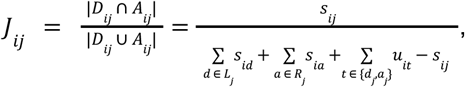

where *s*_*ij*_ denotes the count of split-reads mapping to intron *j* in sample *i, d*_*j*_ is the donor site of intron *j, a*_*j*_ is the acceptor site of intron *j, L*_*j*_ is the the set of introns using *d*_*j*_, *R*_*j*_ is the set of introns using *a*_*j*_, and *u*_*it*_ denotes the count of non-split reads spanning the exon-intron boundary at a splice site *t*.

### Denoising autoencoder

As for the original FRASER, we modeled and controlled for sample covariation using an autoencoder that takes as input a matrix X consisting of the logit-transformed splice metrics of each intron *i* and sample *j* using the following formula:

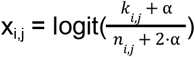

where *k*_*ij*_ and *n*_*ij*_ are the numerator and the denominator of the Intron Jaccard Index of intron *j* in sample *i*, and *α* corresponds to a pseudocount needed to avoid taking the logarithm of or dividing by zero. Originally, in FRASER a pseudocount of 1 was used as a default. Here we assessed various possible pseudocount values.

### Annotation of genes to introns

To report results on the gene level, FRASER uses a user-provided gene annotation file to assign introns to genes. Each intron is assigned to the gene(s) overlapping either the donor or the acceptor site of the intron in a strand-specific manner. As such, an intron may contribute to the gene-level *P*-value of several genes. Genes fully contained in an intron but not overlapping either splice site are not assigned to the intron.

### Multiple testing correction and effect size cutoff

As in FRASER, *P*-values are obtained for each intron by modeling the Intron Jaccard Index using beta-binomial regression on the autoencoder latent space (23). We used the same strategy as in FRASER to obtain gene-level *P*-values. Specifically, intron-level *P*-values are corrected using Holm’s method per gene and sample, and the minimal corrected *P*-value is selected. Then, false discovery rate (FDR) correction of those selected *P*-values is applied across genes per sample using the Benjamini-Yekutieli method. This FDR correction step is by default done transcriptome-wide across all expressed genes. Genes are considered to be expressed if they have at least one intron assigned to them in the dataset. FRASER 2.0 introduces the additional option to restrict the FDR correction to user-provided lists of genes. The provided genes can differ between samples. To define outlier status, the FRASER 2.0 default cutoffs are FDR ≤ 0.1 and effect size *Δ*J ≥ 0.1. FRASER 2.0 additionally introduces an option to flag results located in blacklist regions of the genome (33) as those results may be less trustworthy.

### Rare variant recall benchmarks

To perform rare variant recall analyses, four different sets of splice affecting variants were considered, each defined by a different variant effect prediction tool. The first set contained all splice-site and splice-region variants as defined by VEP (31), corresponding to 1–3 bases of an exon or 1–8 bases of an intron. For the second set, we used SpliceAI (9) and considered variants with a SpliceAI score ≥ 0.5. For the third set, we used MMSplice (7) and considered variants with an |Δ logit *Ψ*| score ≥ 2. The fourth set contained variants predicted to cause aberrant splicing by AbSplice, specifically those with a maximum score across tissues except testis ≥ 0.05, which corresponds to the medium suggested cutoff. Rare variants were defined by a gnomAD MAF < 0.1% and present in at most 2 individuals of GTEx.

In each GTEx tissue, genes were ranked based on the nominal gene-level *P-*value, and the recall of rare splice affecting variants was calculated at each rank. To facilitate comparison across tissues and ensure that the reported outliers are relevant, we used two different measures of performance for each tissue: 1) the recall and precision of ranking the nominal *P*-values of FDR significant results, and 2) the recall at the rank corresponding to a mean of 20 outliers per sample for each tissue.

### Correlation with mapped reads

The number of mapped reads was computed using the function idxstats from SAMtools (32). Then, the spearman correlation coefficient and *P-*value between the mapped reads and its splicing outliers per sample were calculated using the cor.test function in R.

### Tissue reproducibility analysis

To assess the reproducibility of our splicing outlier calls, we called aberrant splicing events across all 48 GTEx tissues both with FRASER and FRASER 2.0, as well as SPOT and LeafcutterMD, using default settings for all methods. This analysis was done on the set of individuals with at least 20 out of the 48 tissues and genes with available *P-*values in at least 25 out of the 48 tissues. This led to 14,707 genes for FRASER 2.0, 16,104 genes for FRASER, 12,154 genes for SPOT and 14,001 genes for LeafcutterMD to be considered. For all methods, we applied three nominal *P-*value cutoffs of 10^−5^, 10^−7^ and 10^−9^ and computed for each gene-sample combination that passes this cutoff in at least one tissue in how many other tissues it could be reproduced with a nominal *P-*value < 10^−3^.

### Benchmark on previously reported pathogenic splice events

For the Yépez et al. dataset (*N*=303), 26 cases were found to have a splice defect in the disease causal gene with FRASER. Out of those 26 cases, 4 harbored large deletions which were excluded from the benchmark, resulting in a set of 22 cases with validated pathogenic splice defects.

## RESULTS

### Introduction of the Intron Jaccard Index as a robust splice metric

To be less sensitive to local splice site aberrations that do not have a strong effect on the canonical splice isoform, we introduce the Intron Jaccard Index, a new intron-centric metric that integrates alternative donor usage, alternative acceptor usage and intron retention signal (Figure 1B). We define the Intron Jaccard Index *J*_*ij*_ as the Jaccard Index of the set of donor-associated reads *D*_*ij*_ and the set of acceptor-associated reads *A*_*ij*_ for a given sample *i* and intron *j*:

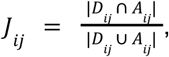

where *D*_*ij*_ contains, for sample *i*, both the split-reads with the same donor site as intron *j* and the non-split reads spanning the exon-intron boundary at that donor site and analogously for *A*_*ij*_ (Figure 1B, Materials and methods). The intersection of those two sets represents the split-reads mapping exactly to intron *j* while the union captures all reads mapping to any intron with the same donor or acceptor site as intron *j* in addition to non-split reads at those splice sites.

The inclusion of both split and non-split reads in the metric allows retaining FRASER’s capability to capture several types of aberrant splicing, including partial or full intron retention, within a single metric (Figure 1C). As in FRASER, we model the Intron Jaccard Index with a beta-binomial denoising autoencoder and calculate *P-*values with the beta-binomial distribution to assess the statistical significance of each Intron Jaccard Index value. Outliers are then called based on the multiple testing corrected *P-*values and effect size (ΔJ).

We ran FRASER with the Intron Jaccard Index as the splice metric on several GTEx tissues. Following the rationale that rare variants in the vicinity of splice sites are likely to disrupt splicing (33), we evaluated our new metric on the recall of rare (MAF < 0.1%) splice-disrupting candidate variants using a combination of variant annotation tools (Materials and methods). On this benchmark, adopting the Intron Jaccard Index metric increased the recall compared to the three metrics from FRASER together and individually (Figure 1D, Supplementary Figure 1).

### Optimization of FRASER 2.0 parameters

After having established the Intron Jaccard Index, we evaluated several FRASER parameters to identify their optimal values with respect to the new metric. As FRASER’s autoencoder works with values in the logit space, which is defined for values greater than 0 and less than 1, a pseudocount needs to be added to both the numerator and denominator when calculating each metric on raw read counts. So far we had set the pseudocount to 1. Here we investigated a range of possible values and found that reducing the pseudocount improved the recall of rare splice-disrupting candidate variants across 15 GTEx tissues, with the optimum being reached at 0.1 (Supplementary Figure 2A,B). We further investigated the effect size cutoff. The highest recall of splice-disrupting candidate variants was achieved for a difference between the observed and expected value, denoted ΔJ, of 0.2 consistently across different filtering settings (Supplementary Figure 3). However, the more permissive cutoff ΔJ = 0.1 was similarly optimal. We therefore decided to adopt the Δ = 0.1 cutoff as the default (Supplementary Figure 2C,D).

Furthermore, we investigated the intron filtering criteria. FRASER uses three parameters, denoted *k, n*, and *q* to filter out lowly expressed introns. The parameter *k* is the minimal value of the splice metric numerator required in at least one sample. The parameter *n* is the minimal value of the splice metric denominator at percentile *q* across samples. Among parameter values with optimal performance, we opted for the most permissive setting (*k*=20, *q*=25% and *n*=10, Supplementary Figure 2E). This parameter setting means that to be considered by the algorithm in a first place, an intron must be supported by at least 20 split reads in at least one sample (Intron Jaccard Index numerator) and shall have a total of more than 10 donor-site and acceptor-site related reads (Intron Jaccard Index denominator, Figure 1B) in at least 25% of the samples. Adopting these cutoffs resulted in testing 287,943 introns and 16,548 coding and non-coding genes on average for each GTEx tissue (Supplementary Figure 2F).

Finally, we investigated whether discarding outlier calls in introns with a poor goodness-of-fit would improve the performance further. Generally, a poor fit can indicate a violation of the modeling assumption, making the *P-*value estimates not trustworthy. FRASER models the data of each individual intron using a beta-binomial regression on the latent space. We observed some instances for which the modeling assumption was violated, in particular cases of splicing quantitative trait loci (sQTL) *in cis* that were not captured by the latent space (Supplementary Figure 4C,D). To systematically capture those cases, we considered the beta-binomial overdispersion parameter (*ϱ*) as a measure of the goodness-of-fit. However, filtering out introns with a high overdispersion parameter had minimal effects on the overall recall of rare variants. Therefore, we decided to not implement this filtering (Supplementary Figure 4A,B).

Overall, we call this new approach FRASER 2.0. All the parameters for which we explored the optimal default values here can be changed by the user.

### Improved performance and robustness of FRASER 2.0 on GTEx

We ran FRASER 2.0 on 48 GTEx (V8) tissues and benchmarked it against FRASER (23), LeafcutterMD (21) and SPOT (22). Notable sample-sample correlations were present in raw Intron Jaccard Index values analogous to what has been previously described in FRASER and the autoencoder of FRASER 2.0 was able to correct for them (Supplementary Figure 5). Introns that were detected as outliers in FRASER but not with FRASER 2.0 tended to have a high *Ψ*_5_ or *Ψ*_3_ value and a low Intron Jaccard Index value, whereas the splice metric values of introns that were found as outliers by both methods were similar (Supplementary Figure 6). FRASER 2.0 *P-*values were better calibrated than FRASER’s *P-*values for each metric (Figure 2A, Supplementary Figure 7), confirming that the Intron Jaccard Index metric is more robust.

**Figure 2.**
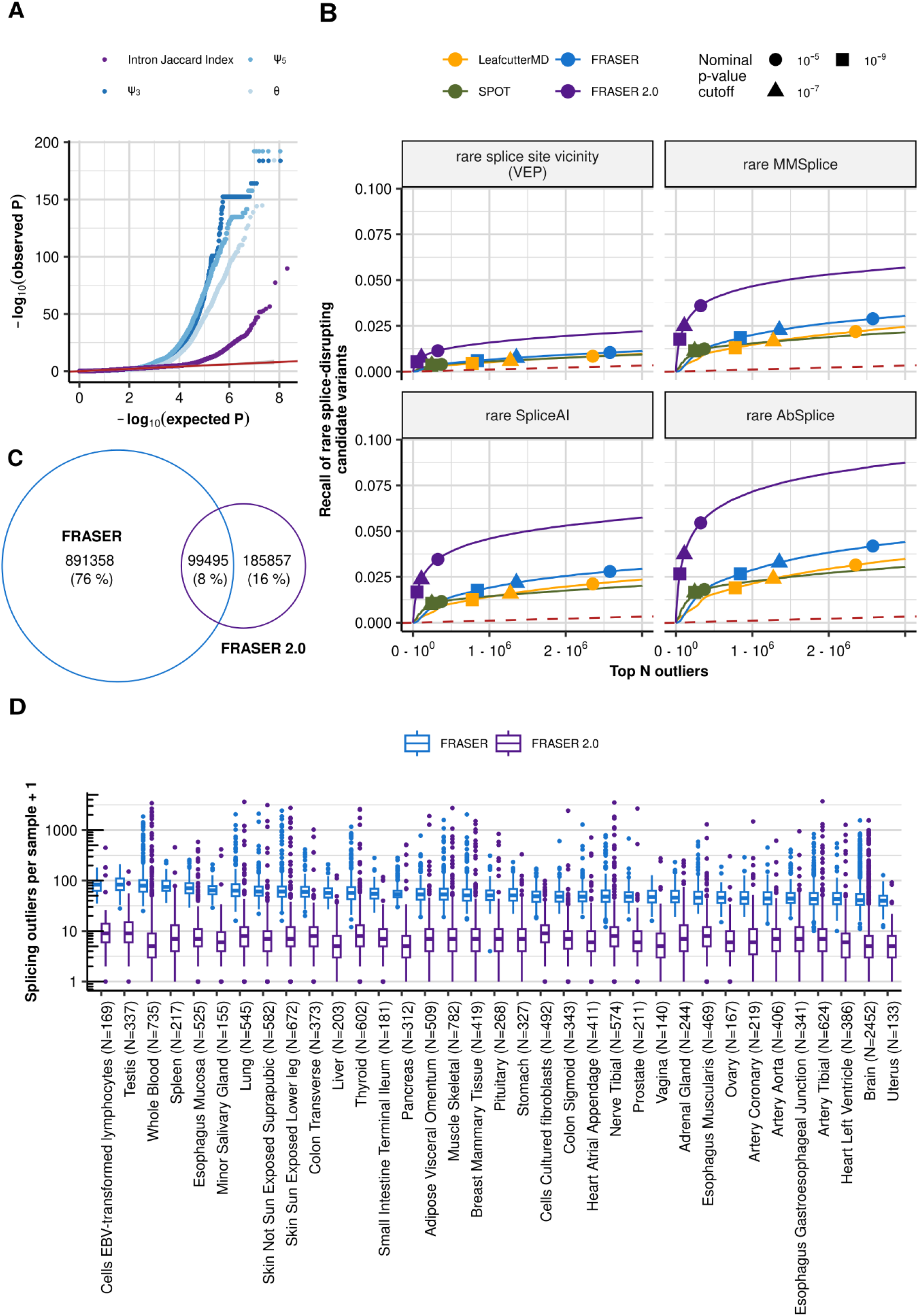
FRASER 2.0 increases recall of rare splice-disrupting candidate variants on GTEx. **(A)** Quantile-quantile plots of the *P-*values for the different splice metrics from FRASER (*ψ*_3_, *ψ*_5_, *θ*, shown in shades of blue) and the Intron Jaccard Index metric from FRASER 2.0 (purple) on the GTEx skin not sun exposed dataset. The red line depicts the diagonal and the gray ribbon around it the 95% confidence interval. **(B)** Recall of rare splice-disrupting candidate variants as defined by the variant annotation tools VEP, MMSplice, SpliceAI, and AbSplice (facets) versus the rank of nominal *P-*values combined across GTEx tissues for FRASER (blue), FRASER 2.0 (purple), LeafcutterMD (yellow), and SPOT (green). Nominal *P-*value cutoffs are indicated with shapes. **(C)** Venn diagram of the overlap of splicing outliers at the gene-level found with FRASER (blue) and FRASER 2.0 (purple). **(D)** Boxplots of the number of splicing outliers (gene-level) per sample (y-axis) called by FRASER (blue) and FRASER 2.0 (purple) for each GTEx tissue (x-axis). All brain tissues have been combined for readability.

We then evaluated each method on their ability to identify splicing outliers in genes predicted to be affected by a rare splice-disrupting candidate variant. FRASER 2.0 consistently outperformed FRASER, SPOT and LeafCutterMD and increased the recall of such genes throughout all ranks and across tissues (10 fold increase in precision at FDR=0.1, Figure 2B and Supplementary Figure 8). Importantly, FRASER 2.0 reported significantly less outliers per sample than FRASER on all tissues (10.0 ± 2.6 times less, Figure 2C,D). While the overlap of outliers from FRASER and FRASER 2.0 is only 8% across all tissues (Figure 2C), outliers identified by FRASER 2.0 only were more enriched for candidate rare splice-disrupting variants than those identified by FRASER only (Supplementary Figure 9), highlighting that FRASER 2.0 allows removing outlier calls from FRASER that are not biologically meaningful.

In addition, splicing outliers from FRASER, LeafcutterMD, and SPOT were more likely to arise with a higher sequencing depth than ones detected with FRASER 2.0 (Figure 3), thus confirming the robustness of the latter. FRASER 2.0 outliers were also more often reproducible across tissues than competitor methods (Supplementary Figure 10).

**Figure 3.**
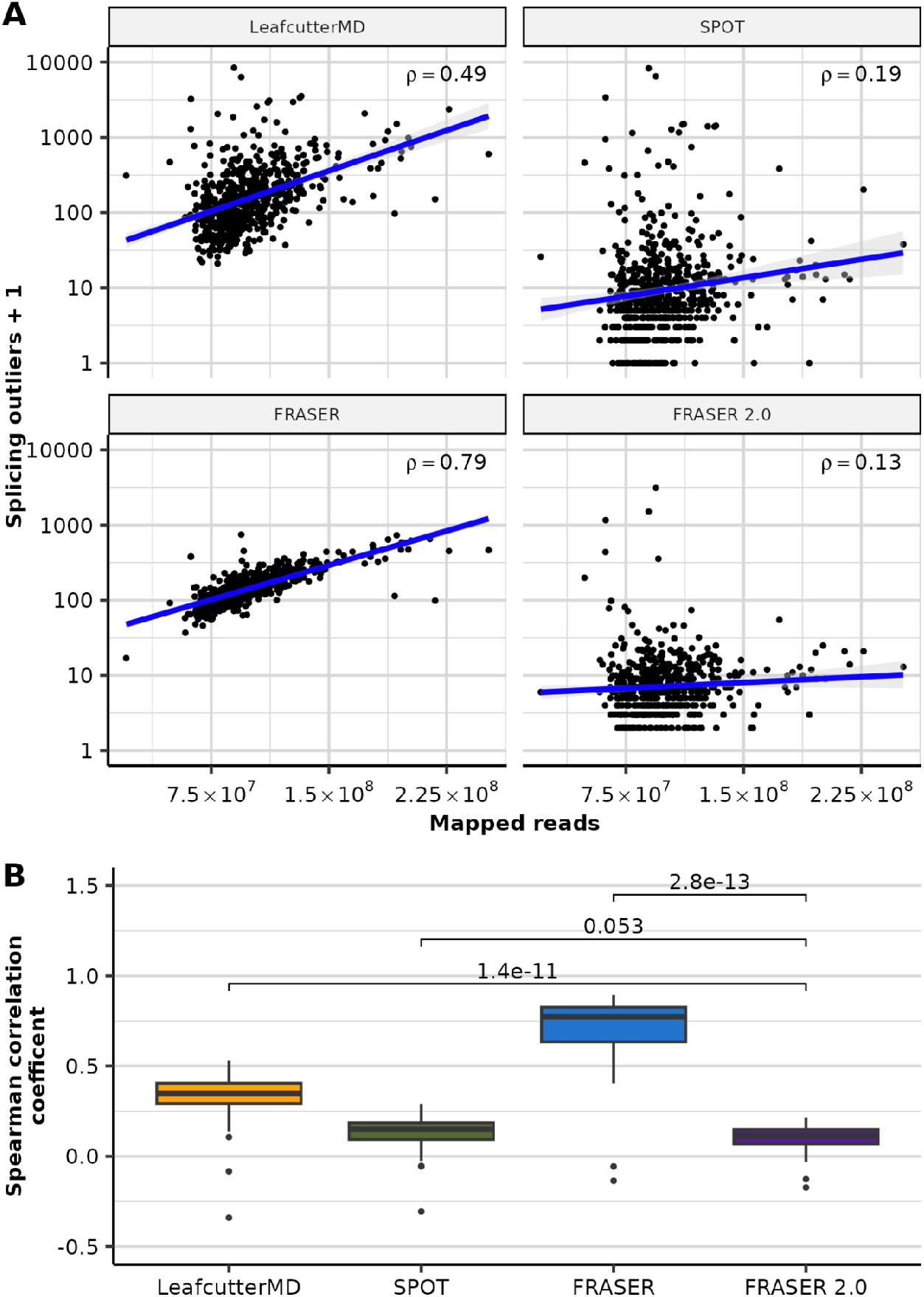
FRASER 2.0 is less sensitive to sequencing depth than previous methods. **(A)** Scatterplot of the number of splicing outliers at the gene level against the total mapped reads per sample on the GTEx skin not-sun-exposed dataset for LeafcutterMD, SPOT, FRASER and FRASER 2.0 (facets). Spearman correlation coefficients (*rho*) are shown. All are significant (Spearman test, *P* < 3 × 10^−3^) **(B)** Boxplots of the Spearman correlation coefficients (y-axis) between the mapped reads and the number of splicing outliers at the gene level called by LeafcutterMD, SPOT, FRASER and FRASER 2.0 (x-axis) for each GTEx tissue (*N*=48). *P*-values of Wilcoxon tests are shown above brackets.

Overall, FRASER 2.0 reports less, but more biologically-relevant splicing outliers than FRASER as well as LeafcutterMD and SPOT.

### Application of FRASER 2.0 to rare disease cohorts

To evaluate FRASER 2.0 in a diagnostic setting, we applied it to 747 samples from the Undiagnosed Disease Network (28) and to the cohort composed of RNA-seq samples from 303 individuals suspected to be affected with a mitochondrial Mendelian disorder (20). As with GTEx, FRASER2 reported less splicing outliers than FRASER in all cohorts (3 to 9.5 times less outliers on median, Figure 4A).

**Figure 4.**
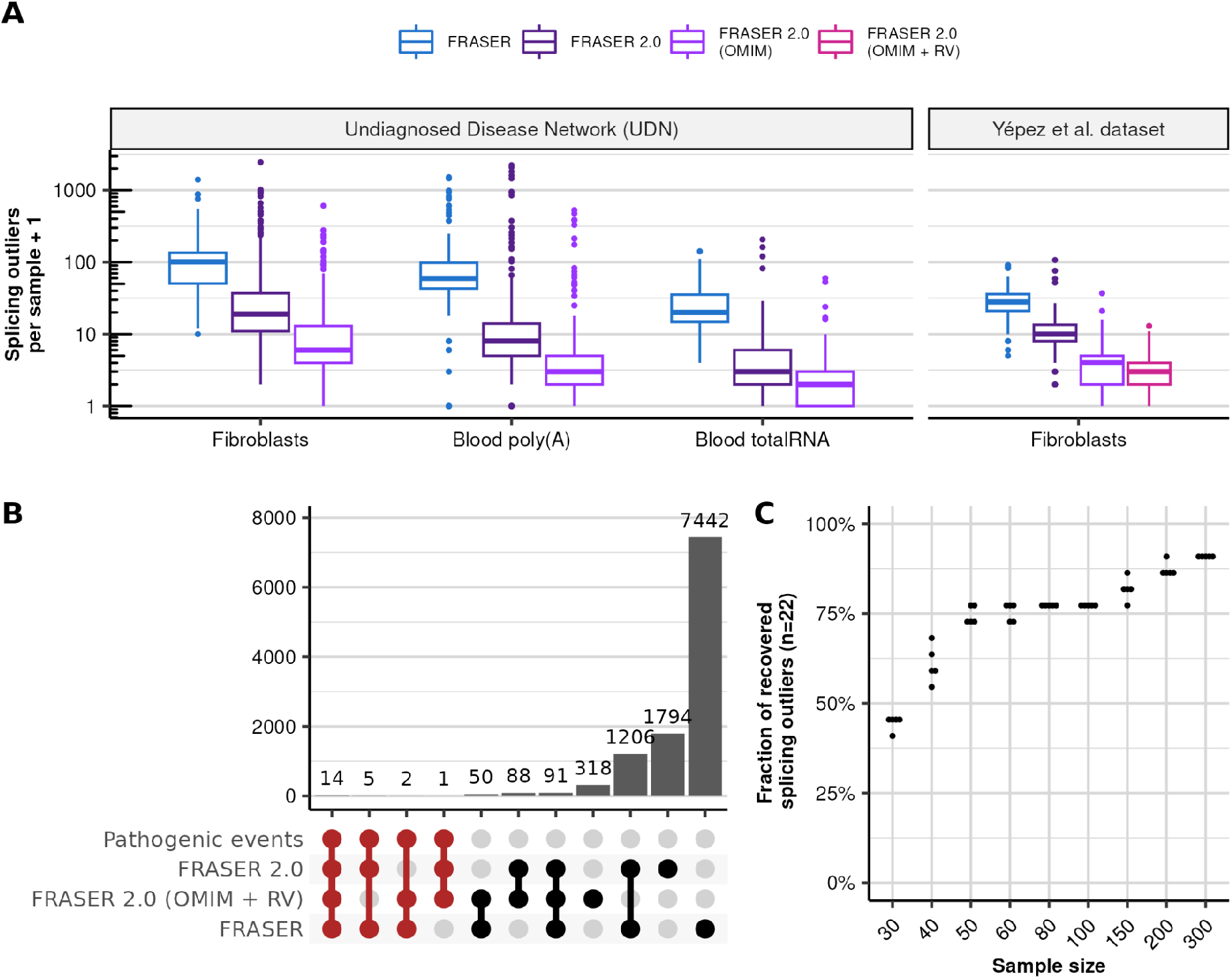
Application of FRASER 2.0 to rare disease cohorts. **(A)** Distribution of the splicing outliers per sample at the gene level on the UDN (*N*=391, *N*=252, *N*=104 for Fibroblasts, Blood poly(A) and Blood totalRNA) and the Yépez et al. dataset (*N*=303) for FRASER (blue) and FRASER 2.0 applied to three gene sets considered for FDR-correction: expressed genes (dark purple), expressed OMIM genes (light purple), and expressed OMIM genes with a rare variant (violet red, Methods). **(B)** Size (bars) of all non-empty intersections (linked dots) between four outlier sets from the Yépez et al dataset: i) the 22 originally reported pathogenic events, ii) the transcriptome-wide significant FRASER 2.0 calls, iii) the significant FRASER 2.0 calls when only considering OMIM genes with a rare variant and iv) the transcriptome-wide significant FRASER calls. **(C)** Fraction of recovered pathogenic splicing outliers from the Yépez et al. dataset (y-axis, total *N*=22) when subsampling to different sample sizes (x-axis). Each sample size was randomly sampled 5 times. RV: rare variant.

Frequently in diagnostics, researchers are not interested in testing all genes, but only those that could cause the disease (e.g. OMIM or curated lists for each disease). In addition, interpretation of variants revealed by panel, whole exome (WES) or whole genome sequencing (WGS) is often performed prior to RNA sequencing, yielding a restricted list of candidate genes per sample. Such prior information has the potential to reduce the multiple testing burden when analyzing splicing outlier calls from RNA-seq. To implement this strategy, we extended the FDR correction step of FRASER 2.0 to allow considering a subset of genes specific for each sample. Testing OMIM genes on both cohorts and OMIM genes harboring at least one rare variant (gnomAD MAF < 0.1%, in cohort allele frequency < 1%, within the gene body except for the UTRs) called by WES in the Yépez et al. dataset consistently led to less outliers per sample to be manually inspected (median of 2 outliers per sample, Figure 4A). The latter approach resulted in a median of 5,427 introns and 149 OMIM genes to be tested per sample, compared to 140,230 introns and 16,846 genes in the transcriptome-wide setting (Supplementary Figure 11).

In the Yépez et al. dataset, 22 cases with aberrant splicing on the disease causal gene were identified by FRASER. FRASER 2.0 reported 20 out of those 22 diagnosed cases with the default cutoffs (Figure 4B). The two missing cases were both exon truncations caused by synonymous variants and showed a large deviation from canonical splicing (ΔJ = -0.55 and ΔJ = -0.39), which was reflected in low nominal *P-*values (1.08 × 10^−4^ and 1.87 × 10^−5^), but due to the multiple-testing burden from testing all expressed introns and genes, the resulting FDR was 1 in both cases. Testing only OMIM genes with rare variants recovered these 2 cases at 10% FDR (FDR = 0.087 and 0.094). However, 5 other cases were not detected with this approach because the gene affected by the splice defect was not in the list of tested genes. In one case, the causal variant had a MAF of 0.0096 which is above the 0.1% cutoff we adopted here, and the 4 other cases had splicing defects caused by intronic variants that were not detected by WES (Figure 4B). Calling variants from WGS could help to overcome this issue and further improve the usefulness of this feature.

Finally, we investigated the sensitivity of FRASER 2.0 to sample size as the number of samples is often limited in rare disease cohorts. We used the Yépez et al. dataset and the 22 known pathogenic splicing events to estimate the required dataset size to reach significance for most of the clinically relevant events. As expected, the percentage of recovered pathogenic events dropped with reduced sample size (Figure 4C, Supplementary Figure 12). With already 50 samples, we recovered 78% of cases (17 out of 22, on average).

Altogether, these results show the general applicability of FRASER 2.0 to various RNA-seq cohorts and that it reports considerably less outliers than FRASER. In addition, the new functionality to test a preselected number of genes per sample can further help reduce the search space. FRASER 2.0 is publicly available at github.com/gagneurlab/fraser and integrated into the workflow DROP (30).

## DISCUSSION

Here we introduced FRASER 2.0, a method to detect aberrant splicing using a novel intron-centric metric, the Intron Jaccard Index. In a single metric, the Intron Jaccard Index captures former metrics of splicing efficiency as well as alternative donor and acceptor site choice. The use of a single metric in FRASER 2.0 translates into easier interpretation of results, reduced runtime, computational resources, and storage. Benchmarks on assumed-healthy samples of the multi-tissue dataset GTEx showed that FRASER 2.0 decreases the number of reported splicing outliers by one order of magnitude, recovers splicing outliers associated with candidate splice-disrupting rare variants more accurately than competitor methods, and is more robust to variations in sequencing depth. Application to two unrelated rare disease cohorts further confirmed the relative advantage of FRASER 2.0 over its predecessors.

Motivated by applications with various collaborators, we have here introduced the option to filter genes based on prior information when performing multiple-testing correction. In practice, it is often the case that RNA-sequencing is performed after an inconclusive WES or WGS, which may have yielded variants of unknown significance for which RNA-seq constitutes the needed functional assay. The possibility to focus on candidate genes and variants in such a “DNA-first, RNA-second” mode of operation can be valuable as we have shown on the mitochondrial rare disease dataset application.

Initially, we introduced the Intron Jaccard Index to focus on more functionally relevant outliers than with the original three splicing metrics of FRASER. However, our benchmarks are based on candidate splice-disrupting variants without consideration for canonical splice isoform abundance. Perhaps, these benchmarks, along with a stronger robustness to sequencing depth, show that the main advantage of the Intron Jaccard Index is merely statistical. An explanation for this could be that the Intron Jaccard Index may be more stable as its denominator accounts for a larger set of reads compared to the three splicing metrics of FRASER.

One limitation of FRASER 2.0 and other methods designed to detect outliers is that a sufficiently large cohort is needed. In the rare disease field, attaining these minimal requirements can be especially challenging. Integrating unaffected samples or samples from individuals suffering from other disorders can help overcome this. In addition, count matrices are provided for a variety of tissues in DROP which can be downloaded and integrated with the local samples. In any case, the aggregated samples should have been probed from the tissue and sequenced using a similar protocol. Another limitation of calling aberrant splicing in RNA-seq data is that the gene might not be sufficiently expressed in the probed tissue, and the choice of tissue is usually limited to clinically-accessible ones (e.g. blood or skin). FRASER2 output, however, could be used as an input on the recently-developed AbSplice-RNA method (12) to predict aberrant splicing in multiple tissues.

Another limitation of this study is that we used the GTEx dataset for tuning FRASER 2.0’s parameters as well as comparison to competitor methods. However, even the version of FRASER 2.0 without optimized parameters clearly outperformed previous methods (Figure 1D). Moreover, due to insufficient available data, we have not assessed in other datasets whether the parameters fitted on GTEx remain optimal on other datasets. As we share our analysis pipeline, users with a large enough cohort could reconduct the parameter optimization on their own dataset.

Long-read sequencing shall eventually lead to more direct measurements of isoform expression and to more accurate quantifications of the expressed isoforms, especially at complex loci (34). Such direct estimations of the abundance of functional splice isoforms on every locus hold the promise to advantageously substitute local splicing metrics including the intronic Jaccard index. However, because of the higher cost and limited sequencing depth of long-read sequencing, which is particularly limiting for RNA-seq due to the high dynamic range of gene expression, short-read based methods such as FRASER 2.0 will probably remain relevant for rare-disease diagnostics in the coming years.

In conclusion, FRASER 2.0 can be readily used in the context of rare disease diagnostics as it has the same sensitivity of FRASER and reports less, but more relevant outliers thus facilitating manual candidate gene inspection. We anticipate the option to include prior knowledge and allow the restriction of the FDR correction to sets of candidate genes to be especially useful in diagnostic strategies where DNA sequencing is routinely done as a first step and RNA-seq is used as a follow-up. As splicing-based therapeutics are on the rise (3, 35), we hope FRASER 2.0 will be a useful tool for the identification of the exact aberrant splicing event for these new targeted therapies.

## Data Availability

No new data was generated for this study. The GTEx data used in this manuscript was obtained from dbGaP accession number phs000424.v8.p1. The intron counts of the Yépez et al. dataset were downloaded from Zenodo. The UDN dataset was obtained from dbGaP accession number phs001232.v4.p2.

https://doi.org/10.5281/zenodo.4646823

https://doi.org/10.5281/zenodo.4646827

https://www.ncbi.nlm.nih.gov/projects/gap/cgi-bin/study.cgi?study_id=phs001232.v4.p2

https://gtexportal.org/home/protectedDataAccess

## AVAILABILITY

No new data was generated for this study. For a description of the used datasets, refer to Materials and Methods. FRASER 2.0 is an open-source R/Bioconductor package available in the GitHub repository https://github.com/gagneurlab/fraser (version 1.99.0 and above). It is also integrated into the workflow DROP (version 1.3.0 and above) available at https://github.com/gagneurlab/drop. The code to reproduce the figures in this manuscript can be found at https://github.com/gagneurlab/FRASER-2.0-analysis.

## SUPPLEMENTARY DATA

Supplementary Data are available at NAR online.

## AUTHOR CONTRIBUTION

Conceptualization: I.S., C.M., V.A.Y., J.G.; Methodology: I.S., C.M., V.A.Y., J.G.; Software: I.S.; Validation: I.S., K.L., V.A.Y.; Formal analysis: I.S., K.L., V.A.Y.; Data curation: I.S., C.M., V.A.Y.; Writing - original draft: I.S., V.A.Y., J.G.; Writing - review & editing: all authors; Visualization: I.S., C.M., V.A.Y., J.G.; Supervision: C.M., V.A.Y., J.G.; Project administration: V.A.Y., J.G.; Funding acquisition: J.G.

## ACKNOWLEDGMENT

We would like to thank Nicholas H. Smith and Felix Brechtmann for valuable advice throughout the project, as well as the different people that have tested and used the software.

The Genotype-Tissue Expression (GTEx) project was supported by the Common Fund of the Office of the Director of the National Institutes of Health and by the National Cancer Institute, National Human Genome Research Institute, National Heart, Lung, and Blood Institute, National Institute on Drug Abuse, National Institute of Mental Health, and National Institute of Neurological Disorders and Stroke. The GTEx data used for the analyses described in this manuscript were obtained from dbGaP accession number phs000424.v8.p1. The Undiagnosed Diseases Network (UDN) is supported in part by the Intramural Research Program of the National Human Genome Research Institute and by grants U01HG007674, U01HG007703, U01HG007709, U01HG007672, U01HG007690, U01HG007708, U01HG007530, U01HG007942, U01HG007943, U01TR002471, U54NS093793, U54NS108251, U01HG010215, U01HG010233, U01HG010217, U01HG010230, U01HG010219, and U01TR001395 from the National Institutes of Health Common Fund, through the Office of Strategic Coordination/Office of the NIH Director. The content is solely the responsibility of the authors and does not necessarily represent the official views of UDN investigators or the National Institutes of Health. The datasets used for the analyses described in this manuscript were obtained from dbGaP at http://www.ncbi.nlm.nih.gov/gap through dbGaP accession number phs001232.

