## Supplementary Figures for "Improved detection of aberrant splicing using the Intron Jaccard Index"

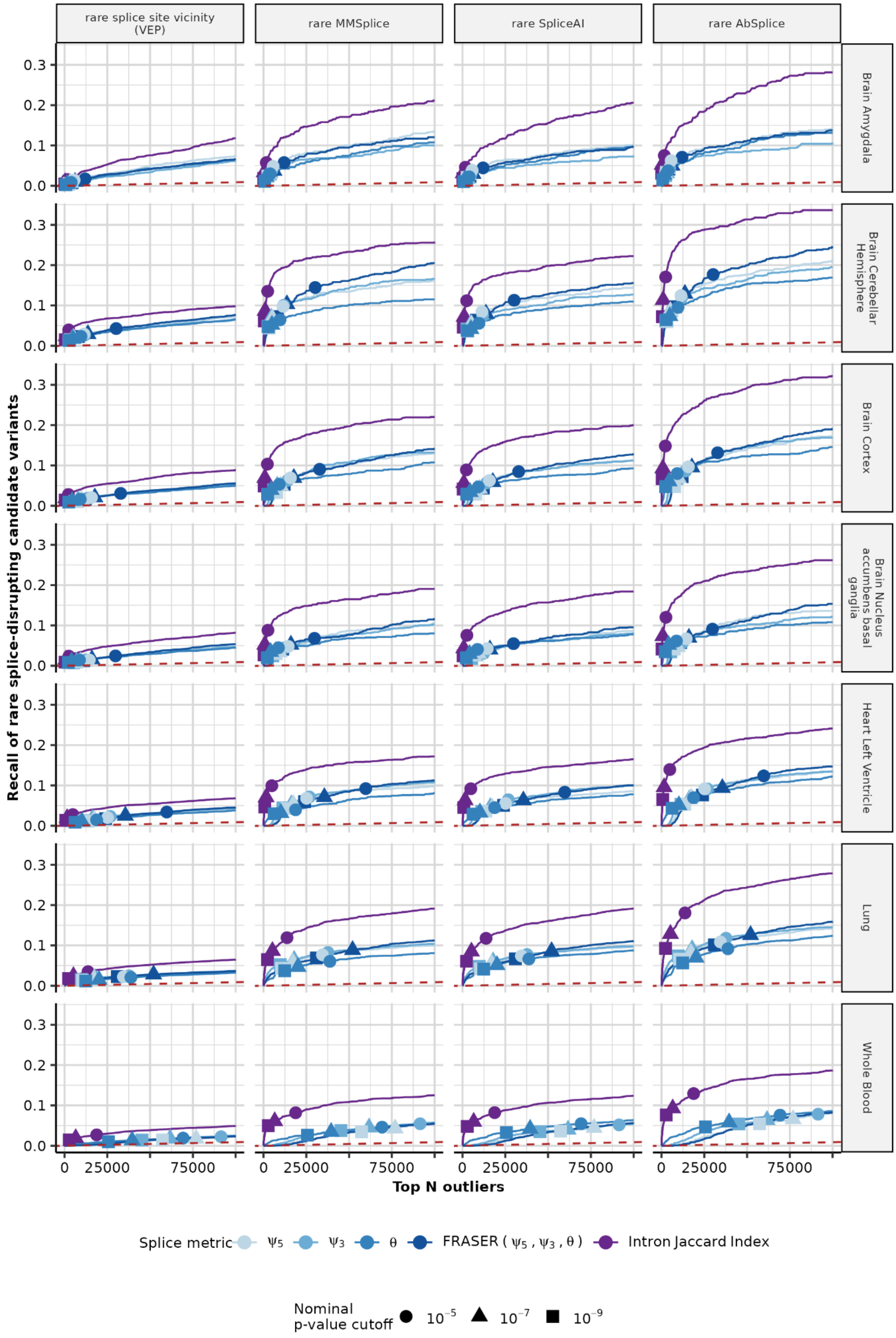

rare splice site vicinity (VEP)
rare MMSplice
rare SpliceAI
rare AbSplice
Brain Amygdala
Brain Cerebellar Hemisphere
Brain Cortex
Brain Nucleus accumbens basal ganglia
Heart Left Ventricle
Lung
Whole Blood
Recall of rare splice-disrupting candidate variants
0.0
0.1
0.2
0.3
0
25000
75000
Top N outliers
Splice metric
ψ5
ψ3
θ
FRASER ( ψ5 , ψ3 , θ )
Intron Jaccard Index
Nominal p-value cutoff
10^-5
10^-7
10^-9

**Figure S1: Intron Jaccard Index increases recall of splice-disrupting candidate variants over FRASER's splice metrics on several GTEx tissues.**

Same as Fig. 1D), but for the FRASER adaption using Intron Jaccard Index (purple) compared to individual and combined metrics of FRASER ($\psi_3$, $\psi_5$, $\theta$) on several GTEx tissues (rows) and four different sets of rare splice-disrupting candidate variants (columns).

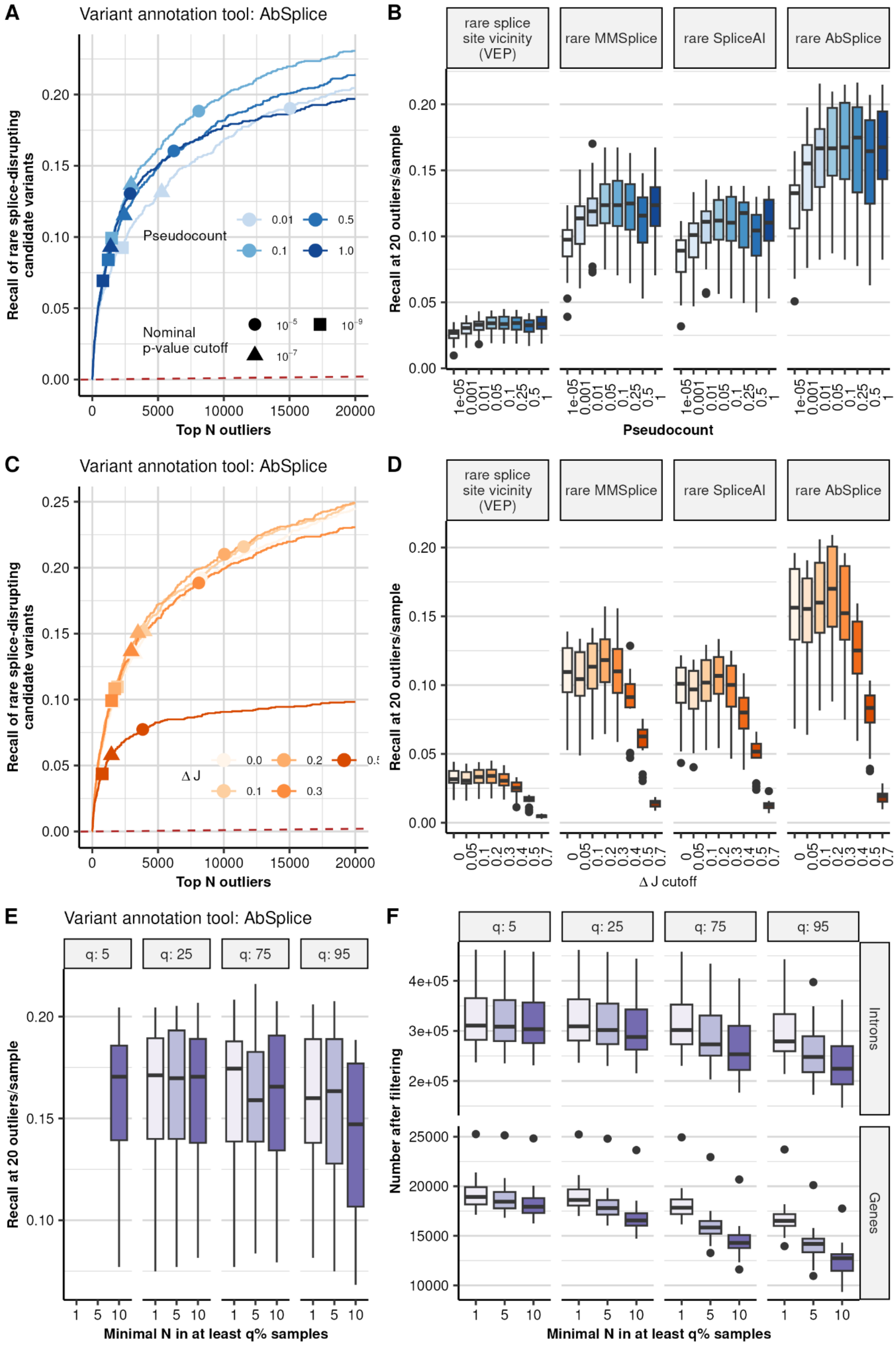
A
Variant annotation tool: AbSplice
Recall of rare splice-disrupting candidate variants
Pseudocount
0.01
0.5
0.1
1.0
Nominal p-value cutoff
$10^{-5}$
$10^{-9}$
$10^{-7}$
Top N outliers
B
rare splice site vicinity (VEP)
rare MMSplice
rare SpliceAI
rare AbSplice
Recall at 20 outliers/sample
Pseudocount
C
Variant annotation tool: AbSplice
Recall of rare splice-disrupting candidate variants
Δ J
0.0
0.2
0.1
0.3
Top N outliers
D
rare splice site vicinity (VEP)
rare MMSplice
rare SpliceAI
rare AbSplice
Recall at 20 outliers/sample
Δ J cutoff
E
Variant annotation tool: AbSplice
q: 5
q: 25
q: 75
q: 95
Recall at 20 outliers/sample
Minimal N in at least q% samples
F
q: 5
q: 25
q: 75
q: 95
Introns
Genes
Number after filtering
Minimal N in at least q% samples

**Figure S2: Identification of optimal FRASER 2.0 parameters pseudocount, Δ jaccard cutoff and filtering settings. (A)** Recall of rare splice-disrupting candidate variants as defined by AbSplice versus the rank of nominal *P*-values from FRASER with the Intron Jaccard Index metric using different values of the pseudocount on the GTEx skin not-sun-exposed dataset. Nominal *P*-value cutoffs are indicated with shapes. **(B)** Boxplots of the recall of rare splice-disrupting candidate variants at the rank corresponding to a value of 20 outliers per sample for different values of the pseudocount across 48 GTEx tissues. Facets indicate different tools to define the set of candidate splice-disrupting variants: VEP (annotated as splice donor/acceptor or splice region), MMSplice (absolute MMSplice Δlogit $\Psi \geq 2$), SpliceAI (SpliceAI score ≥ 0.5) and AbSplice (max. AbSplice score ≥ 0.05). **(C)** Same as (A) but comparing different cutoff values on the predicted Δ Intron Jaccard Index for FRASER with the Intron Jaccard Index metric with pseudocount set to 0.1. **(D)** Same as (B) but comparing different cutoff values on the predicted Δ Intron Jaccard Index for FRASER with the Intron Jaccard Index metric with pseudocount set to 0.1. **(E)** Same as (B) but comparing different filtering settings. Facets indicate the quantile and x-axis indicate the minimal value of N at this quantile to pass the filter. **(F)** Boxplots of the number of introns (top row) and genes (bottom row) after applying the respective filtering setting defined by the quantile (columns) and the minimal value of N at this quantile (x-axis). Boxplots: center line = median; box limits = first and third quartiles; whiskers span all data within 1.5 interquartile ranges of the lower and upper quartiles (applicable here and in all the following figures).

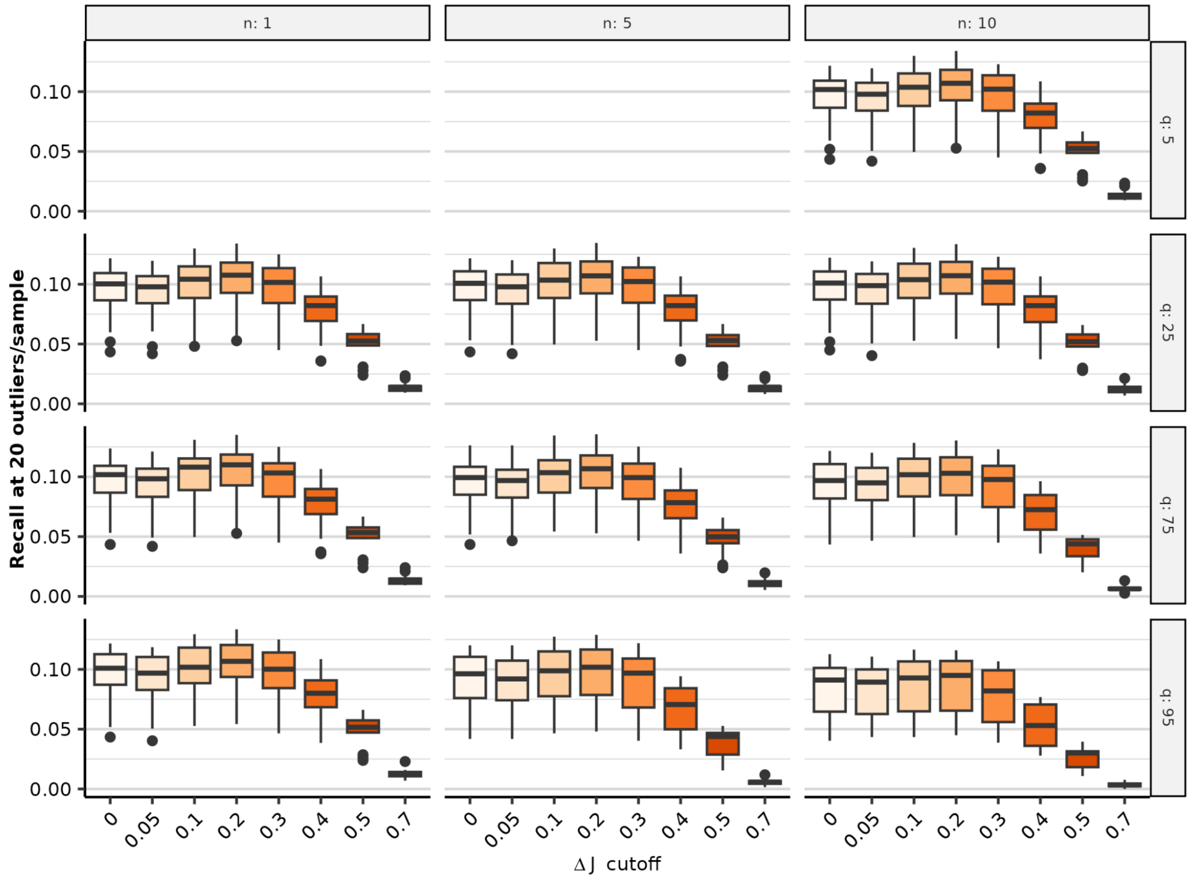


**Figure S3: Combined parameter optimization results for different combinations of ΔJ and filtering cutoffs.**

Boxplots of the recall of rare splice-disrupting candidate variants as predicted with SpliceAI at the rank corresponding to a value of 20 outliers per sample for different values of the ΔJ cutoff across 48 GTEx tissues. Facets indicate different intron filtering settings, defined by the minimal required n (rows) in at least q% of the samples (columns).

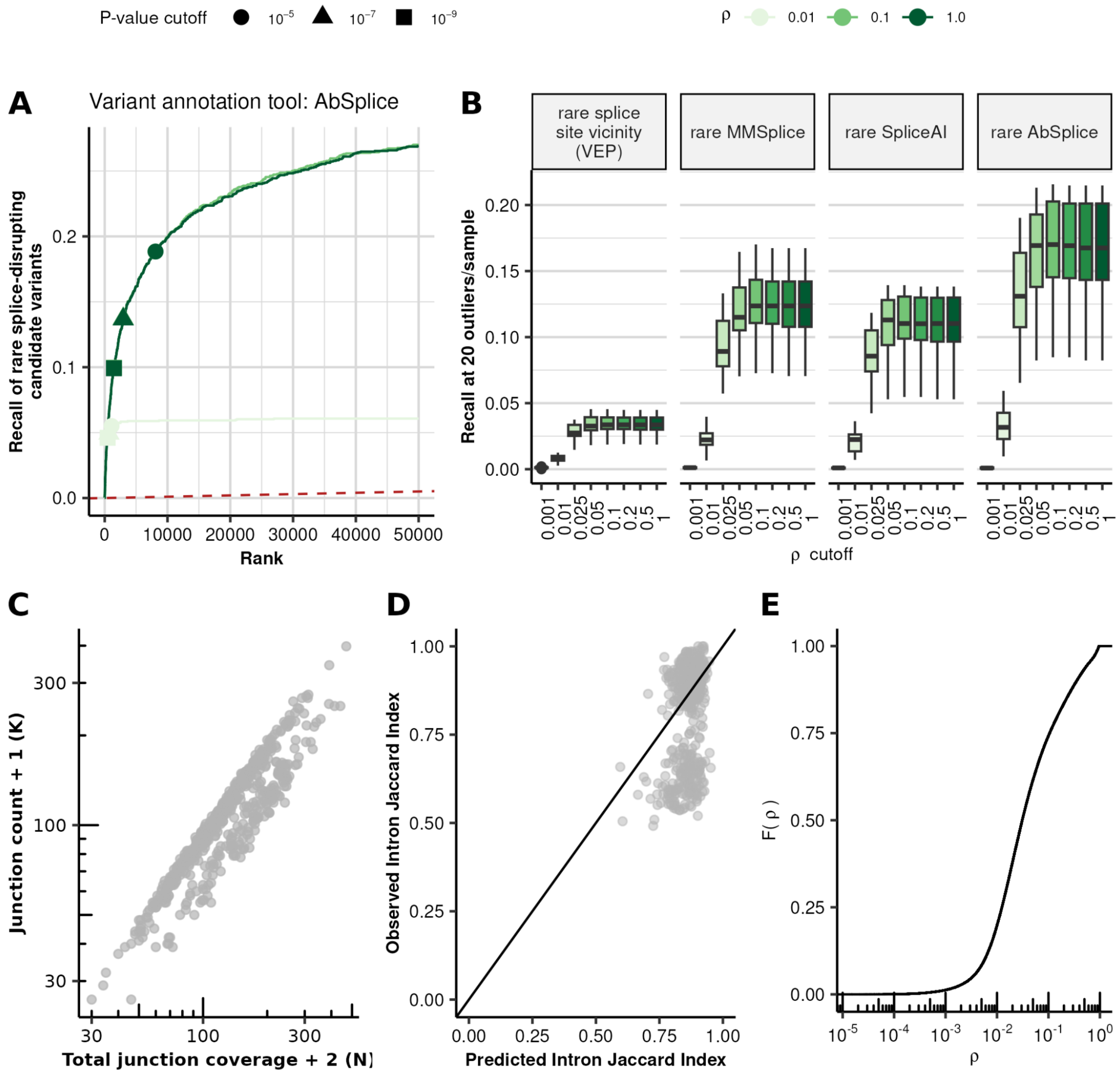


**Figure S4: Goodness-of-fit cutoff does not improve splicing outlier calls. (A)** Recall of rare (MAF < 0.001) splice-disrupting candidate variants as defined by AbSplice versus the rank of nominal *P*-values for FRASER with the Intron Jaccard Index metric and pseudocount of 0.1 for different values of the goodness-of-fit cutoff on $\varrho$ (overdispersion parameter of the beta-binomial distribution, shown in shades of green). Nominal *P*-value cutoffs are indicated with shapes. **(B)** Boxplots of the recall of rare splice-disrupting candidate variants at the rank corresponding to a value of 20 outliers per sample for different values of the goodness-of-fit cutoff across 48 GTEx tissues. Facets indicate different tools to define the set of candidate splice-disrupting variants: VEP (annotated as splice donor/acceptor or splice region), MMSplice (absolute MMSplice Δlogit $\Psi \geq 2$), SpliceAI (SpliceAI score ≥ 0.5) and AbSplice (max. AbSplice score ≥ 0.05). **(C)** Intron counts (y-axis) against the denominator of the jaccard metric (x-axis) of intron chr1:1485839-1486109:+ affected by a sQTL loci in cis across samples in GTEx skin (suprapubic). **(D)** Predicted against observed Intron Jaccard Index values for the intron shown in (C). **(E)** Empirical cumulative density function of $\varrho$ on GTEx suprapubic skin tissue with the Intron Jaccard Index metric and pseudocount of 0.1.

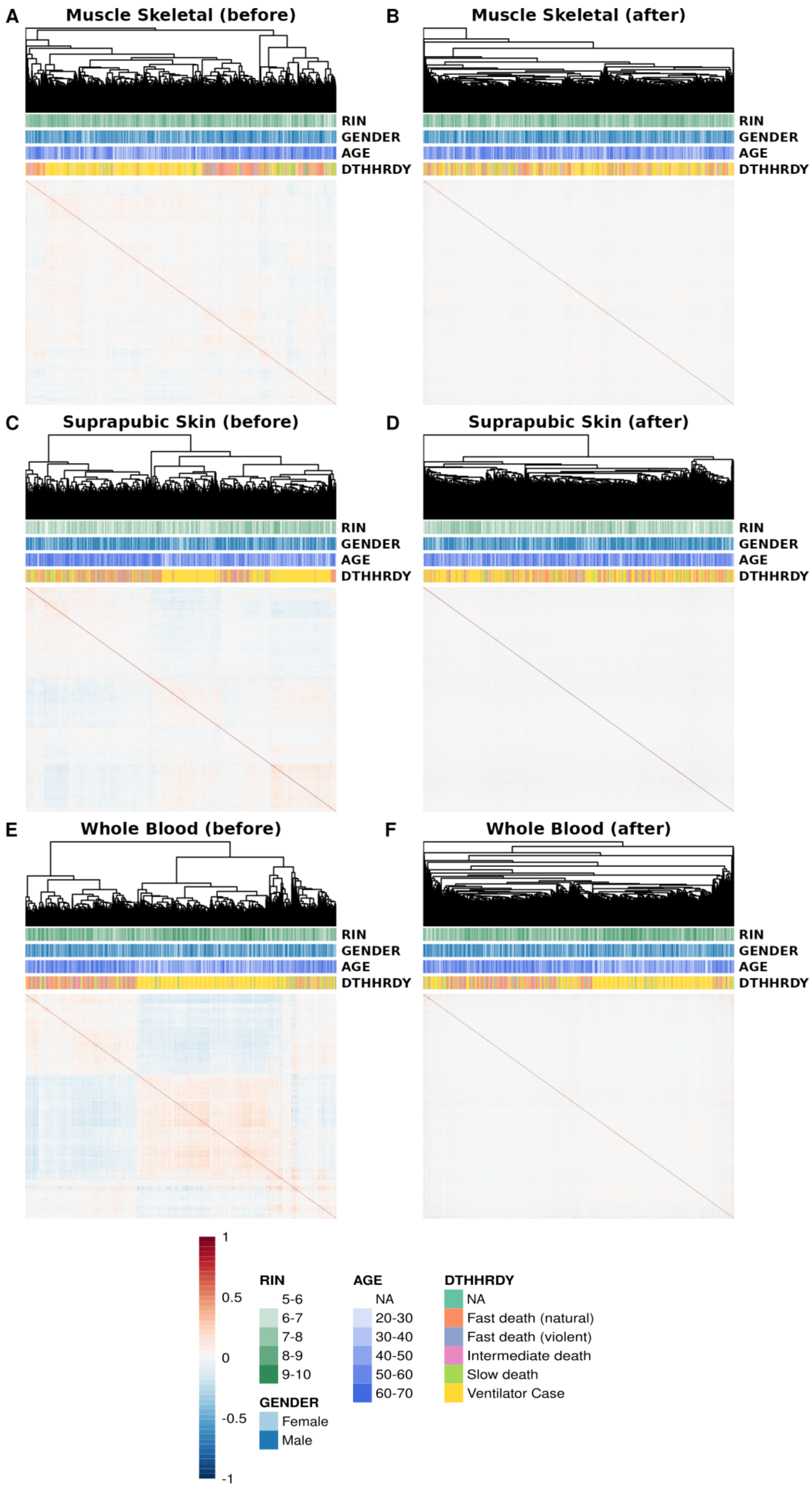
A
Muscle Skeletal (before)
B
Muscle Skeletal (after)
C
Suprapubic Skin (before)
D
Suprapubic Skin (after)
E
Whole Blood (before)
F
Whole Blood (after)
RIN
GENDER
AGE
DTHHRDY
1
0.5
0
-0.5
-1
RIN
5-6
6-7
7-8
8-9
9-10
GENDER
Female
Male
AGE
NA
20-30
30-40
40-50
50-60
60-70
DTHHRDY
NA
Fast death (natural)
Fast death (violent)
Intermediate death
Slow death
Ventilator Case

**Figure S5: Sample correlation heatmaps.** Heatmaps of sample-sample correlations of Intron Jaccard Index metric before **(A, C, E)** and after **(B, D, F)** FRASER 2.0's autoencoder correction on three GTEx tissues: muscle skeletal (**A, B**, *N*=782), not sun-exposed suprapubic skin (**C, D**, *N*=582) and whole blood (**E, F**, *N*=735). A dendrogram of the sample clustering is shown on top of each heatmap alongside sample metadata: RNA integrity number (RIN), age, gender, and cause of death (Hardy scale classification, DTHHRDY).

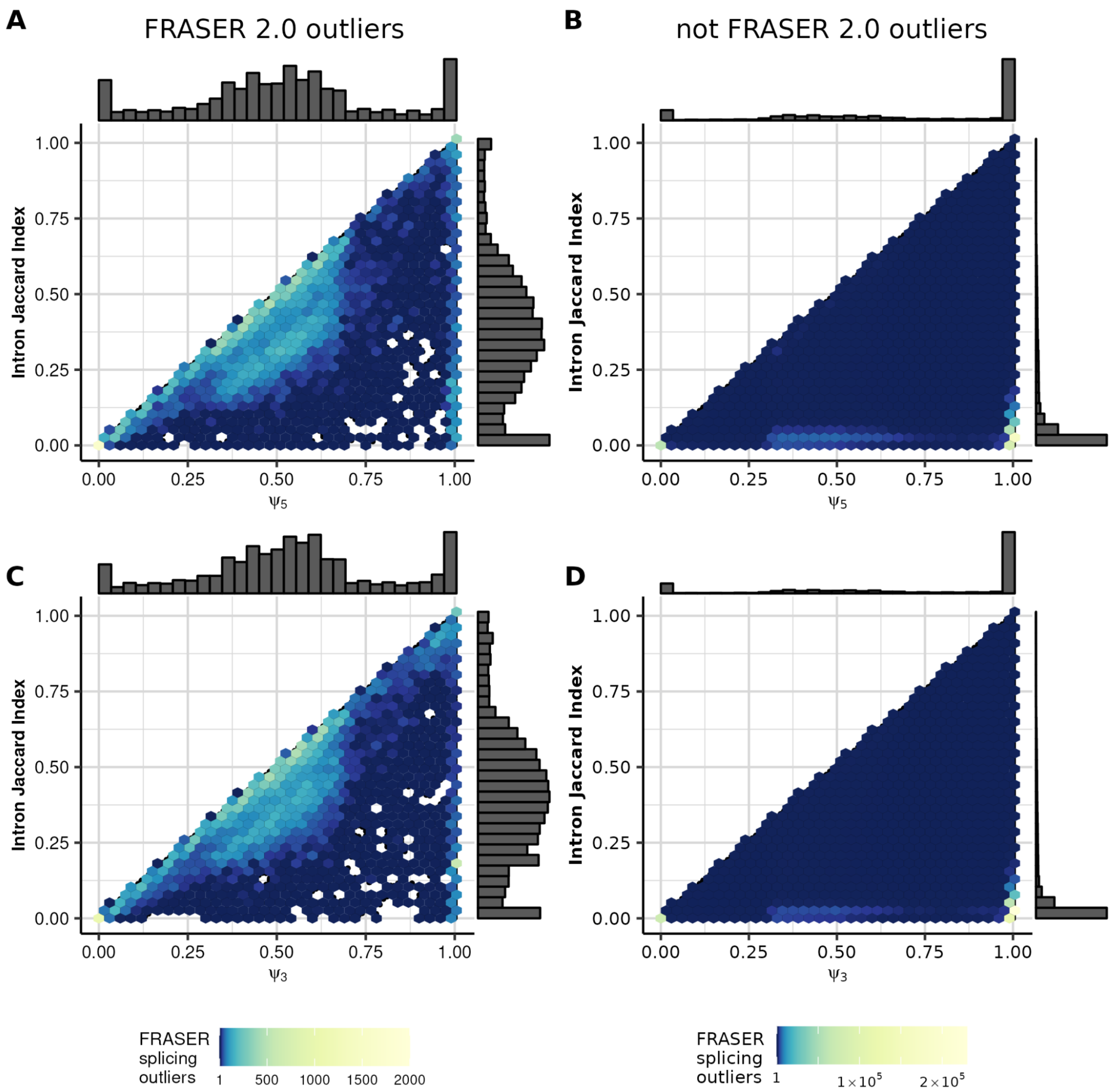


**Figure S6: Comparison of Intron Jaccard Index values to FRASER's $\psi_5$ and $\psi_3$ metrics of FRASER outliers.**

Scatterplot with density of $\psi_5$ values of FRASER $\psi_5$ outliers against Intron Jaccard Index values across GTEx tissues for introns that are also reported as outliers by FRASER 2.0 **(A)** and introns that are not outliers in FRASER 2.0 **(B)**. **(C, D)** Same as (A, B), but for $\psi_3$ values. Outliers that are found both by FRASER and FRASER 2.0 **(A, C)** tend to lie alongside the diagonal, whereas most FRASER outliers that are not reported by FRASER 2.0 **(B, D)** have small values in the Intron Jaccard Index metric while having $\psi_5$ or $\psi_3$ close to 1.

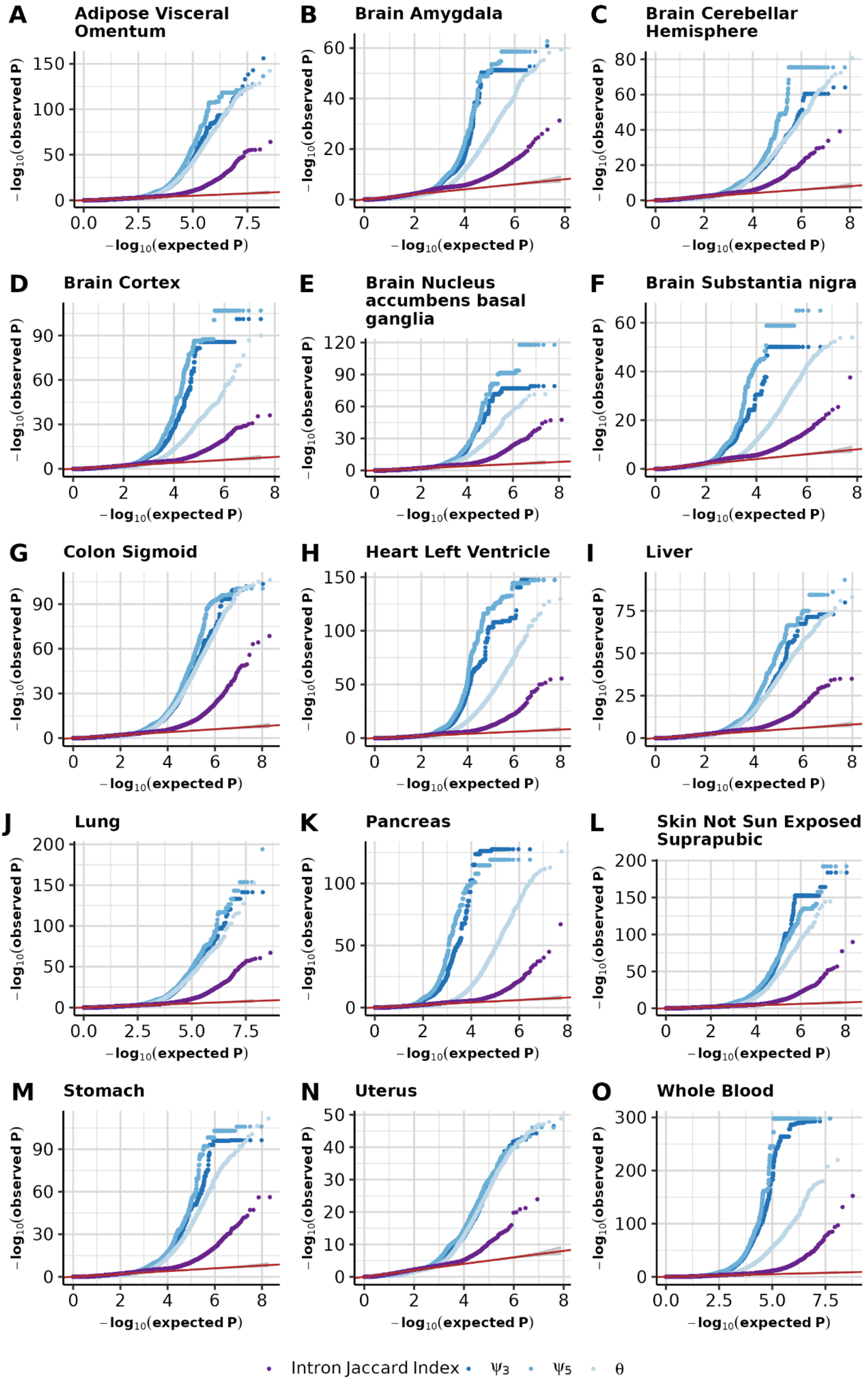

A
Adipose Visceral Omentum
B
Brain Amygdala
C
Brain Cerebellar Hemisphere
D
Brain Cortex
E
Brain Nucleus accumbens basal ganglia
F
Brain Substantia nigra
G
Colon Sigmoid
H
Heart Left Ventricle
I
Liver
J
Lung
K
Pancreas
L
Skin Not Sun Exposed Suprapubic
M
Stomach
N
Uterus
O
Whole Blood
$-\log_{10}$(observed P)
$-\log_{10}$(expected P)
Intron Jaccard Index
ψ3
ψ5
θ

**Figure S7: Quantile-quantile plots of FRASER 2.0 *P*-values.** Quantile-quantile plots of expected against observed *P*-values obtained using the 3 splice metrics from FRASER (different shades of blue) and the Intron Jaccard Index of FRASER 2.0 (purple) on 15 GTEx tissues **(A-O)**. Under the null hypothesis, the data are expected to lie along the diagonal (red, 95% confidence bands in gray).

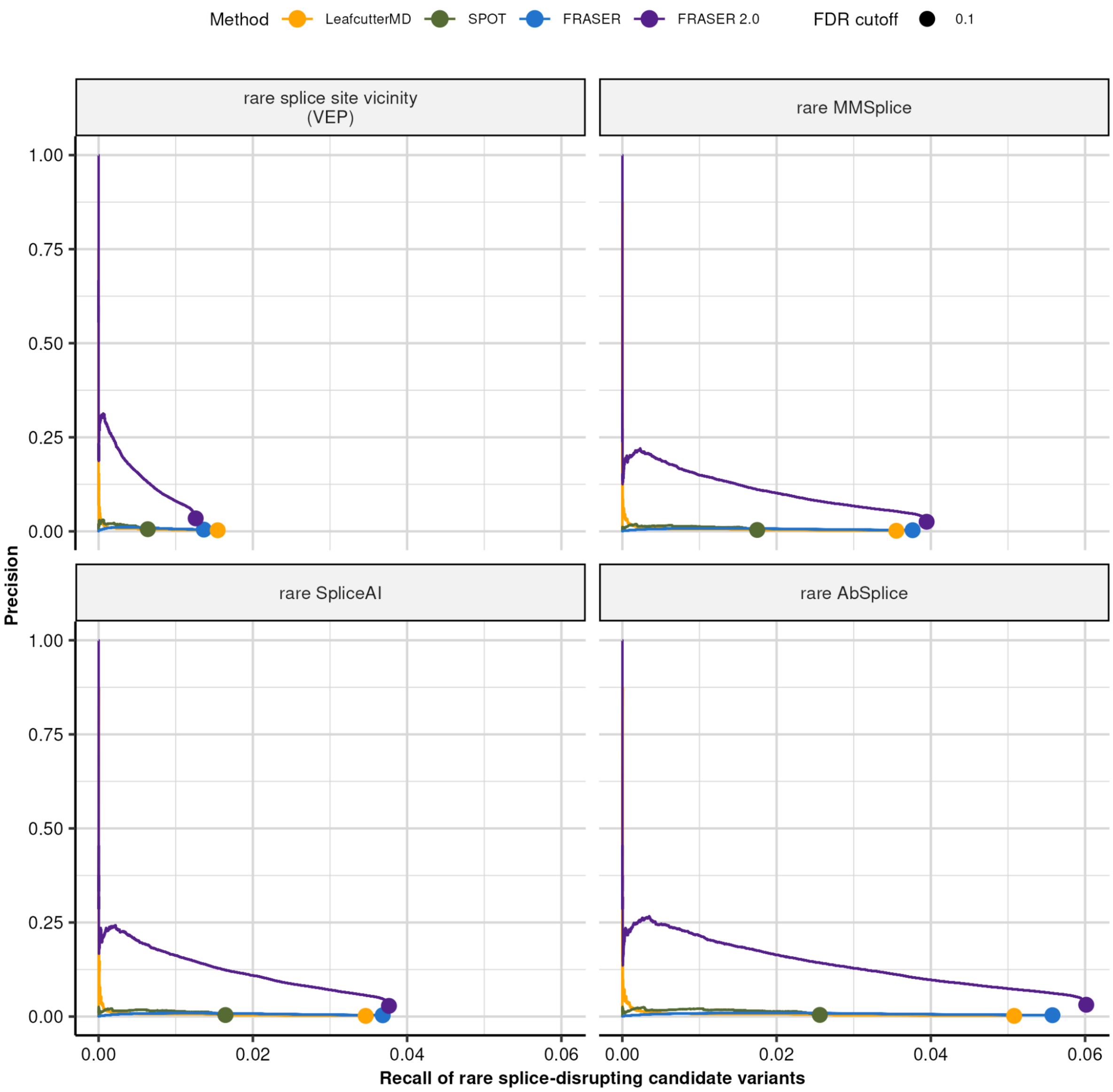


**Figure S8: Improved precision of FRASER 2.0 at FDR cutoff.** Precision-recall plot on candidate rare splice-disrupting variants as defined by the variant annotation tools VEP, MMSplice, SpliceAI, and AbSplice (facets) on nominal P-values combined across GTEx tissues for FRASER (blue), FRASER 2.0 (purple), LeafcutterMD (yellow), and SPOT (green) for significant results at FDR ≤ 0.1. The FDR cutoff is indicated with a circle.

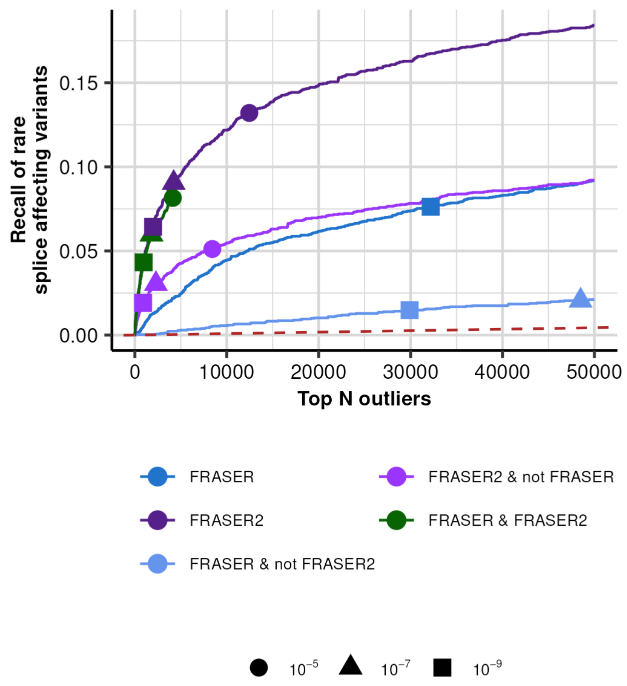


**Figure S9: Recall of splice-disrupting candidate variants by regions of the Venn diagram for FRASER and FRASER 2.0.** Recall of rare splice-disrupting candidate variants as defined by SpliceAI score 0.5 versus the rank of nominal *P*-values combined across GTEx tissues for different regions of the Venn diagram between FRASER (middle blue) and FRASER 2.0 (dark purple) with FRASER-only outliers in light blue, FRASER 2.0 only outliers in light purple and both FRASER and FRASER 2.0 outliers in green. Different nominal *P*-value cutoffs are indicated with shapes.

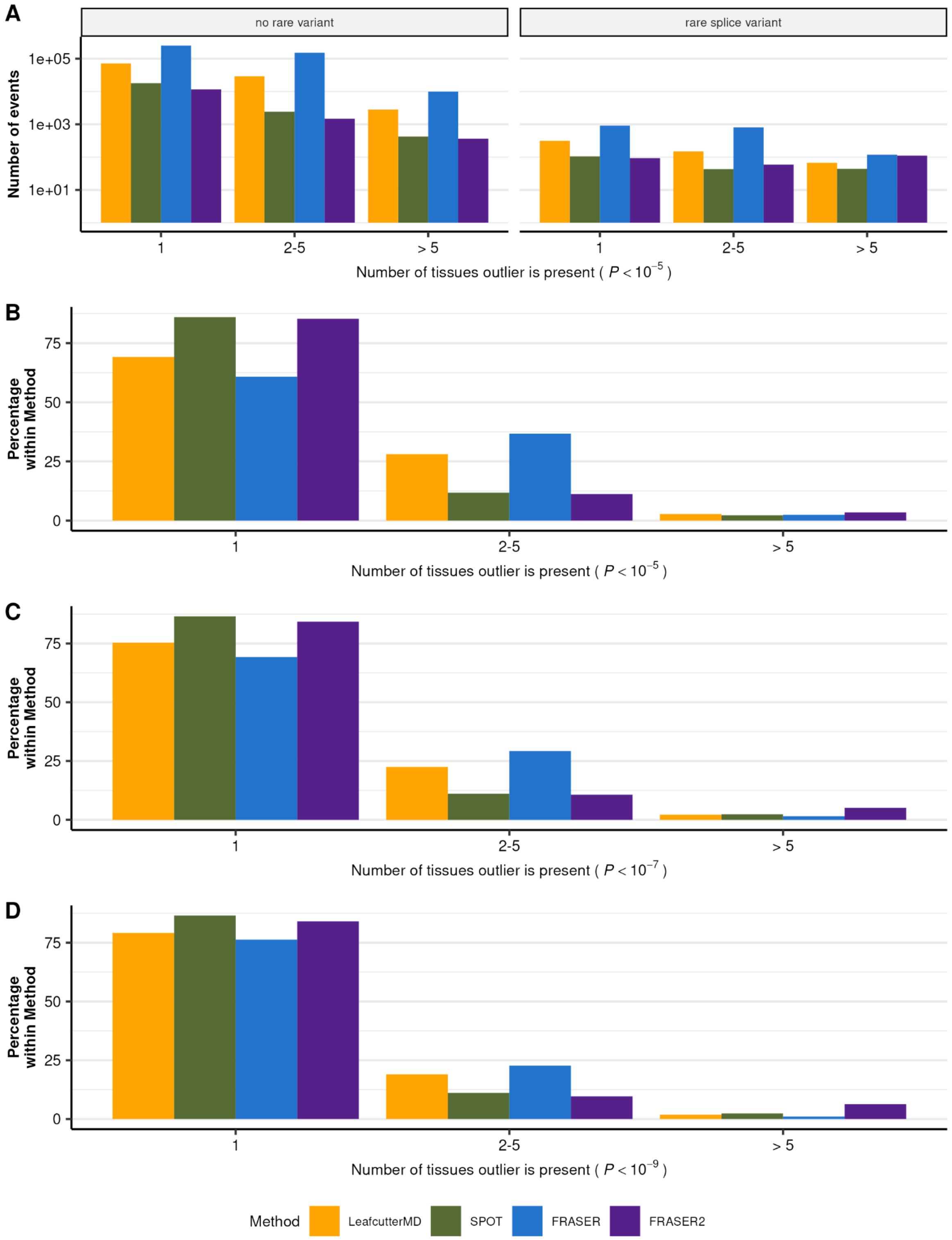


**Figure S10: Reproducibility of splicing outlier calls across GTEx tissues.**
**(A)** Barplot of the number of gene-level splicing outliers (y-axis) against their reproducibility (x-axis) across GTEx tissues. The reproducibility is defined as the number of tissues an event is observed at a nominal *P*-value < $10^{-3}$ given it was observed at least once at a nominal *P*-value < $10^{-5}$. Data is stratified by associated variant status (defined by VEP) and grouped by method. **(B)** Same as (A) but plotted as the proportion (y-axis) of reproducible gene-level splicing outlier calls in GTEx tissues. **(C-D)** Same as (B) but with at least one call at a nominal *P*-value < $10^{-7}$ (C) and nominal *P*-value < $10^{-9}$ (D).

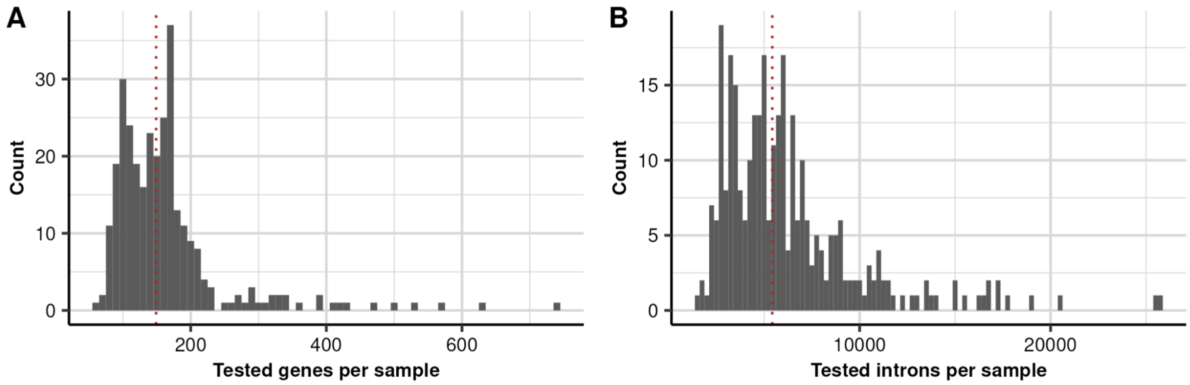


**Figure S11: Tested features per sample on the OMIM + rare variant subset.** Distribution of the total number of tested genes **(A)** and introns **(B)** per sample on the Yépez et al. dataset when including only OMIM genes harboring a rare variant. The red line denotes the median across samples: genes=149 and introns=5,427.

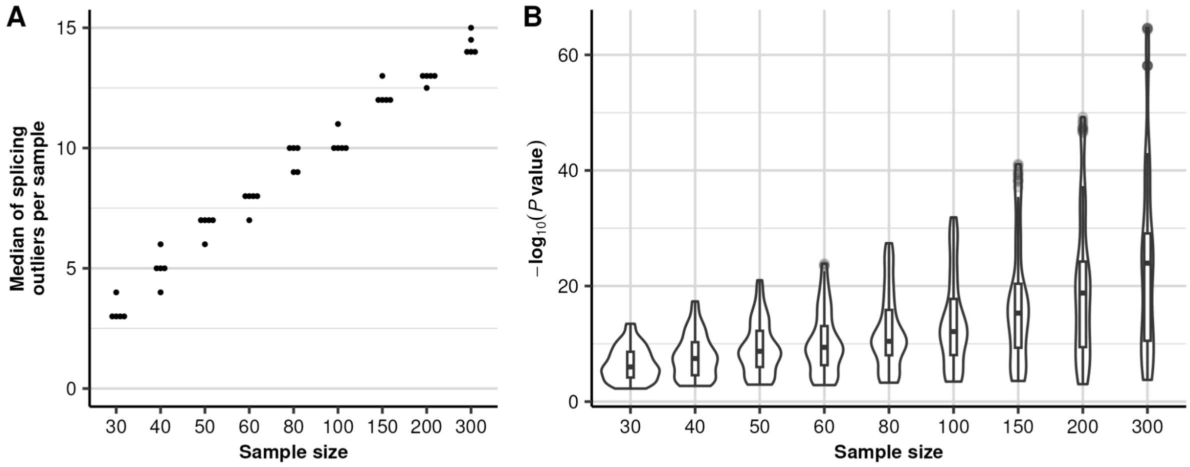


**Figure S12: Power analysis of FRASER 2.0 on the Yépez et al. dataset (*N*=303). (A)** The median of splicing outliers across all samples using default cutoffs (y-axis) is plotted against the taken sample size (x-axis). Each sample size was sampled 5 times. **(B)** The negative $\log_{10}$ *P*-value for all known disease-causing splicing outliers (y-axis) is plotted against the taken sample size (x-axis). The violin depicts the density of the data points.
